# Elevated prevalence and treatment of sleep disorders from 2011 to 2020; a nationwide population-based retrospective cohort study in Korea

**DOI:** 10.1101/2023.05.10.23289759

**Authors:** Eun Kyoung Ahn, Younghwa Baek, Ji-Eun Park, Siwoo Lee, Hee-Jeong Jin

## Abstract

**Objectives:** This study used National Health Insurance claims data from Korea to report the prevalence of sleep disorders and treatment status, including traditional Korean medicine, in the last ten years.

**Methods:** This is a retrospective cohort study in Korea. All diagnosis and prescription data, including herbal medicine claims, from the Health Insurance Review and Assessment Service from 2011 to 2020 were reviewed. Prevalence estimation, direct medical expenses, and prescribed amounts for sleep disorders were recorded.

**Results:** The prevalence of sleep disorders increased from 3,867,975 (7.62%) in 2011 to 7,446,846 (14.41%) in 2020, nearly doubling over 10 years. Insomnia was observed in 91.44% (n=9,011,692) of the patients. The mean number of hospital visits per patient for sleep disorders was 11.5 (±26.62). Benzodiazepines are the most commonly prescribed medications for sleep disorders, and gamma-isoyosan is the most frequently prescribed herbal medicine.

**Conclusions:** Sleep disorders are continuously increasing, as is the use of medical services – personal and social medical expenses are also increasing accordingly. Sleep disorders should be recognized as a significant health problem that needs to be actively addressed to improve quality of life.

**Strengths and limitations of this study:**

- This is a nationwide health insurance claims data for prevalence and the status of treatment on sleep disorders for ten years.
- This study will be meaningful because we have confirmed the current address of the prevalence and treatment of sleep disorders in the last 10 years.
- The data source has the limitation of being able to confirm only the items subject to health insurance benefits and review.
- The results of tests for the diagnosis of sleep disorders were not confirmed.

## INTRODUCTION

Sleep disorders are highly related to chronic pain, cardiovascular disease, dementia, metabolic syndrome, and digestive dysfunction^1-4^, and reportedly increase the death risk, deteriorates quality of life and overall health condition resulting to patient’s economic burden^5,6^. Short-term insomnia occurs in 30–50% of the population^7^. The prevalence of sleep disorders in the United States was 20–41.7% in 2011^8^, and from 2013–2016, the prevalence increased by approximately 40%^9^. The prevalence of insomnia is at least 6% in developed countries^10^, 10% in Norway and England^11,12^, 5.7% in Germany^13^, and 19% in France^14^. In the cases of Asia, Japan recorded 13.5% of insomnia cases in 2016^15^. In Korea, 17–23% of cases were recorded as insomnia, and another study reported 5% of insomnia cases meeting the DSM-IV diagnostic criteria^16,17^. Moreover, in situations of heightened personal and social stress, such as the COVID-19 pandemic, the worldwide prevalence of sleep disorders has surged to approximately 40%^18^. Sleep disorders are becoming common in our society; it is considered that the scale of physical, mental, economic, and social damage is very likely to increase in the long term. Sleep disorders may appear and disappear temporarily but often persist chronically. Reports on the nature of insomnia show that approximately 46% of patients have experienced the symptoms continuously for 3 years^19^, while other studies have shown that approximately 45% of patients continuously experience insomnia after its onset^20,21^.

There are both nonpharmacological and pharmacological treatment strategies for sleep disorders. Typically, pharmacological therapy is provided if there is no response to cognitive behavioural therapy (CBT) or if treatment is not feasible. Medication prescribed for sleep initiation or maintenance disorders is distinguished^22-24^. The European Sleep Research Society (ESRS) treatment guidelines for sleep disorder treatment recommend that benzodiazepines and z-drugs should not be used for long-term treatment^23^. Moreover, the American College of Physicians (ACT) treatment guidelines recommend using these drugs for a short period (4–5 weeks) only, as approved by the US Food and Drug Administration (FDA)^22^. Chronic use of sleeping pills increases the risk of side effects such as daytime sleepiness, ataxia, dizziness, cognitive decline, increased aggressive behavior, delirium, worsening of apnea, and increased risk of dementia^25-28^. In Korea and China, patients may choose sleep disorder treatments such as acupuncture or herbal medicine as alternatives to initial treatment or long-term medication. According to the Clinical Practice Guideline (CPG) of Korean medicine for insomnia disorders, insomnia treatment is divided into herbal medicine, acupuncture, and non-acupuncture herbal medicine treatment^29^, depending on the symptoms, acupuncture, and herbal medicine alone or in combination treatment takes place.

Sleep disorders are gradually increasing in prevalence and they are a health issue requiring long-term approaches. However, no precise treatment has been developed for sleep disorders. Treatment strategies, such as cognitive therapy, acupuncture, and medication, including herbal medicine, are used in clinical practice to treat sleep disorders. Sleep disorder is not a direct condition that causes death in humans; however, it is a condition that affects the quality of life, disease prognosis, and mortality. Hence, it is crucial to grasp the current status of its prevalence and common treatment.

In this study, based on data claims from the Korean National Health Insurance Service, sleep disorders over the past 10 years (2011-2020) included medical and institutional information on both Western and Korean medicine, including covered medications, sleep tests, and treatment prescriptions. This study determined the prevalence and overall treatment characteristics for sleep disorders in Korea.

## METHODS

### Data source and study population

In the current retrospective cohort study, a set of research data (No. M20210819448) was provided by the Healthcare Big Data Hub of the Health Insurance Review and Assessment Service (HIRA) of Korea. The research data were pseudonymized, and provision was determined by the HIRA’s Public Data Provision Deliberation Committee. The study’s protocol was also exempted from ethical review by the Institutional Review Board of the Korea Institute of Oriental Medicine (IRB No. I-2107/006-001).

The data included all patients diagnosed with sleep disorders as a primary or secondary diagnosis between January 1, 2011, and December 31, 2020. Medical records containing information on sleep disorder diagnoses were extracted based on the diagnosis code registered by the treating doctor. Individuals diagnosed with sleep disorders at least once during the observation period and prescribed medication were enrolled in this study. Subjects of all ages were included; however, veterans were excluded from the study.

### Sleep disorders

The Korean Standard Classification of Diseases version 7 (KCD-7) was used to identify subjects with sleep disorders. The KCD-7 code is based on the International Classification of Diseases version 10 (ICD-10) code system and reflects the unique disease characteristics of Korea. The subdivision classification for frequent diseases, reorganization of the Korean medicine classification, and diagnostic codes for rare diseases are also reflected. Sleep disorders were classified according to the criteria of the third edition of the International Classification of Sleep Disorders (ICSD-3)^24^; insomnia, sleep-related breathing disorders, sleep-related movement disorders, circadian rhythm sleep-wake disorders, central disorders of hypersomnolence, parasomnia, and other unspecified sleep disorders. The sleep disorder classification, KCD-7, and detailed diagnoses are provided in Supplementary table 1.

### Medications for sleep disorders

Drugs approved by the Korea Food and Drug Administration (KFDA) for insomnia are classified into benzodiazepines, non-benzodiazepines, antidepressants, antihistamines, and antipsychotics. Korea has a dual medical service system; therefore, herbal medicine prescriptions are available. A list of the licensed drugs for the indication of sleep disorders, including herbal medicines, is provided in Supplementary table 2. In the present study, all 56 types of herbal medicines reimbursed (Supplementary table 3) that are subject to review by HIRA were reviewed. Herbal medicines for sleep disorder indications included Gamisoyo-san, Hwanglyeonhaedok-tang, Galgeunhaegui-tang, Dangguiyukhwang-tang, and Sosiho-tang.

### Data preprocessing and statistical analysis

The data was analyzed by accessing a remote analysis system provided by the Healthcare Big Data Hub. Remote access to a virtualized Personal Computer (PC) is only possible from PCs approved by researchers and authorized in advance. In order to calculate the prevalence of sleep disorders by year, individual subjects diagnosed with sleep disorders and prescribed medications were counted. The total population was calculated based on the population by the end of December of each year, as announced by the Ministry of Public Administration and Security’s Annual Statistical Yearbook Population^25^. It is calculated based on different criteria according to the nature of each variable. Prevalence cases were based on the number of individuals affected with sleep disorders, while the number of cases for medical use was based on the medical use statement. Medication and treatments, on the other hand, were based on individual prescriptions. Age, number of cases of medical use per person, and medical expenses were reported as the mean and standard deviation. The use of Korean and Western medical services, the distinction between hospitalization and outpatient treatment, type of sleep disorder, sex, and medications or test prescriptions were presented as numbers and percentages. Medical expenses were expressed in South Korean Won (KRW). Statistical differences between groups were reported based on a significance level of p <0.05. The Statistical Analysis System (SAS) Enterprise Guide version 9.4.2 (SAS institute Inc., SAS Campus Drive Cary, NC, USA), installed on a virtualized PC, was used for pre-processing and statistical analysis of the research data.

## RESULTS

### General information

A total of 7,467,730 subjects used medical care due to sleep disorder diagnosis, of which 59.62% (n=4,452,628) were female patients, from 2011–2020. The mean age for the entire population was 53.47 ±17.95 years old, while the mean age for men and women was 52.31 ±18.18 and 54.26 ±17.74, respectively. The mean age of women was higher compared to male. When age was divided into 10-year-old units, the number of patients with sleep disorders increased with age, and the highest number of patients was seen in the age group between 50 and 60 years, then gradually decreasing (Table 1). For the distribution by type of sleep disorder according to the ICSD-3, ‘insomnia’ was diagnosed in 91.44% (n= 9,011,692 cases); hence, the most common type of sleep disorder that was recorded in visiting medical institutions. ‘Other unspecified sleep disorders’ accounted for 6.89% (n=679,400), and the rest sleep disorders accounted for less than 1% of total (Figure 1).

**Table 1.**
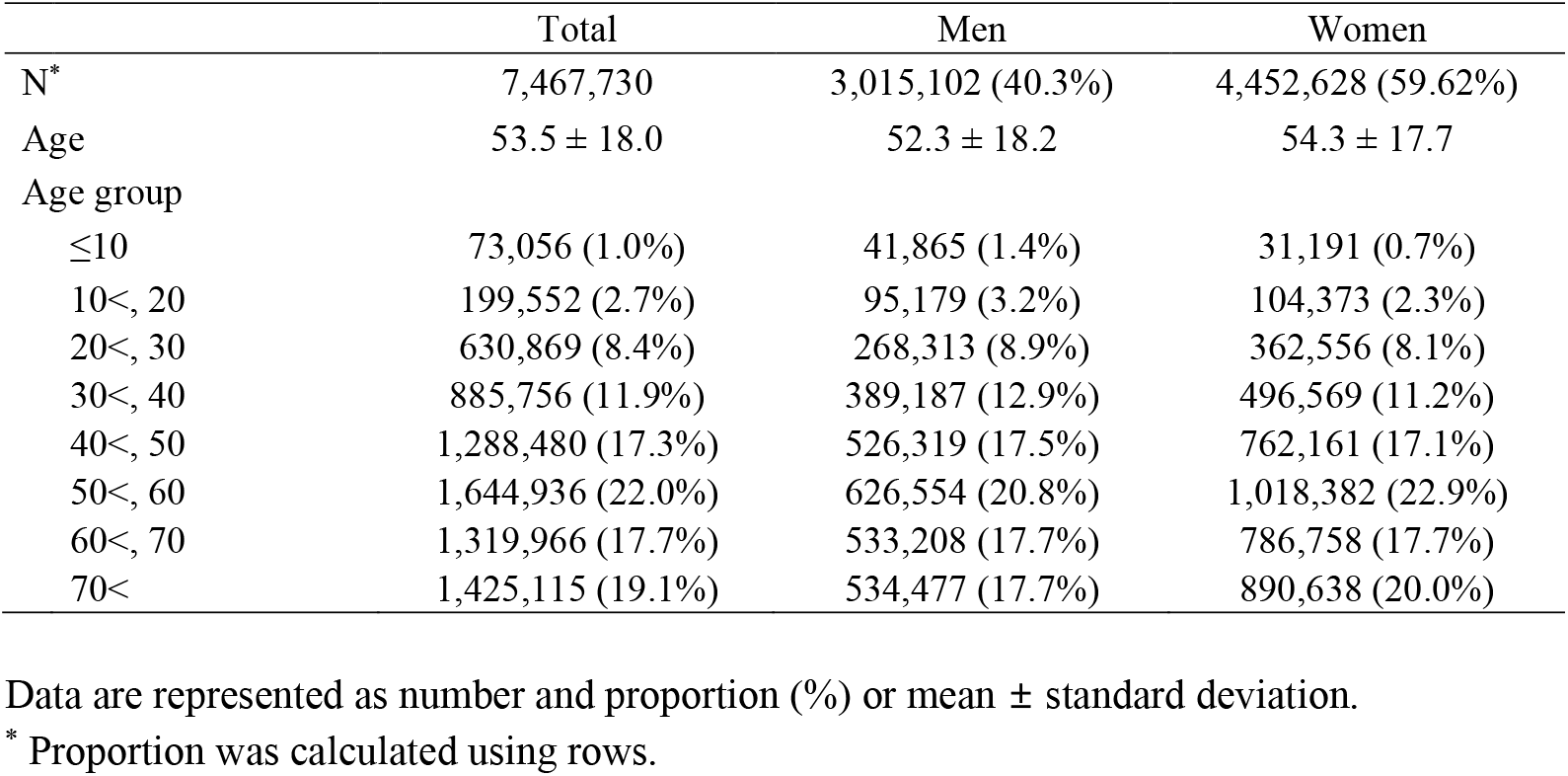
Basic characteristics of study population

**Figure 1.**
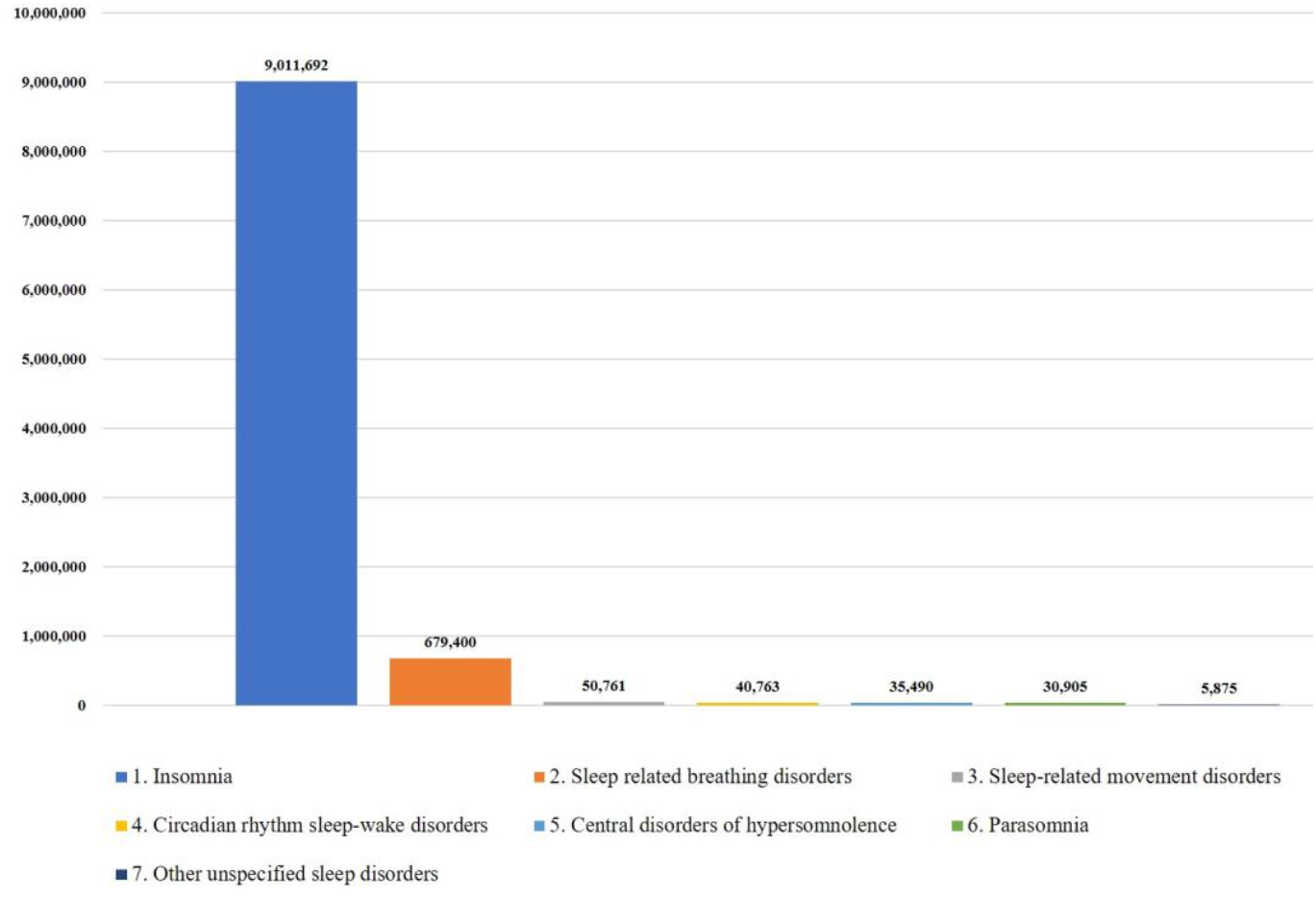
Distribution by type of sleep disorder according to ICSD-3 classification.

### Annual prevalence of sleep disorders and use of medical services

The annual prevalence of sleep disorders in patients aged >10 years increased from 3,867,975 (7.62%) in 2011 to 7,446,846 (14.41%) in 2020, nearly doubling in 10 years. The number of male patients increased from 1,477,614 (5.82%) to 2,987,309 (11.56%), and the number of female patients increased from 2,390,361 (9.44%) to 4,459,537 (17.27%); with rates increase being similar (Figure 2, Supplementary table 4). The mean number of visits to the hospital for sleep disorder diagnosis for the past 10 years was 11.55±26.62, and women visited the hospital (11.96±27.43) more often than men (10.94± 25.37). The total hospital visits also increased from 5, 932, 505 in 2011 to 11,111,518 by 2020. Hospital visits increased from 37.1% to 39.1% in men and decreased from 62.9% to 60.9% among women. There has been an increase in the total number of patients that are women and the number of hospital visits for men (Figure 3). Of the total hospital visits for sleep disorders, Korean medical treatment accounts for approximately 9.65% (N =8,323,747). A total of 11.39% (n=6,066,211) of women visited a Korean medical institute, which was higher than that of men (6.84%, n=2,257,536). The mean number of hospital visits per patient was 11.7±SD 27.4 for women and 10.9±25.4 for men. The mean cost of total medical expenses per visit was 71,931 KRW, and the mean cost per patient was 830,707 KRW (Table 2).

**Figure 2.**
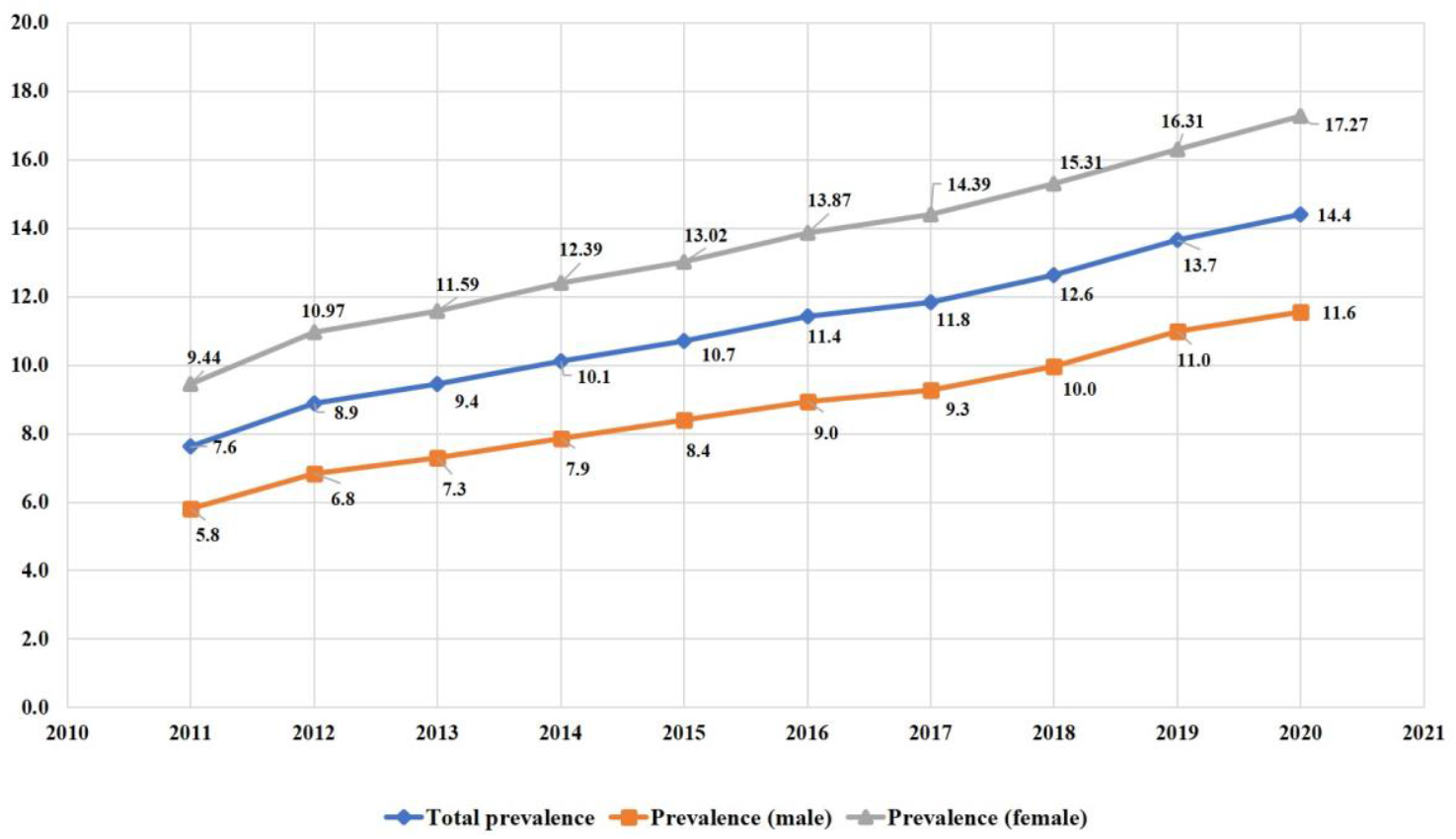
Annual prevalence of sleep disorders for ten years by sex (2011–2020)

**Figure 3.**
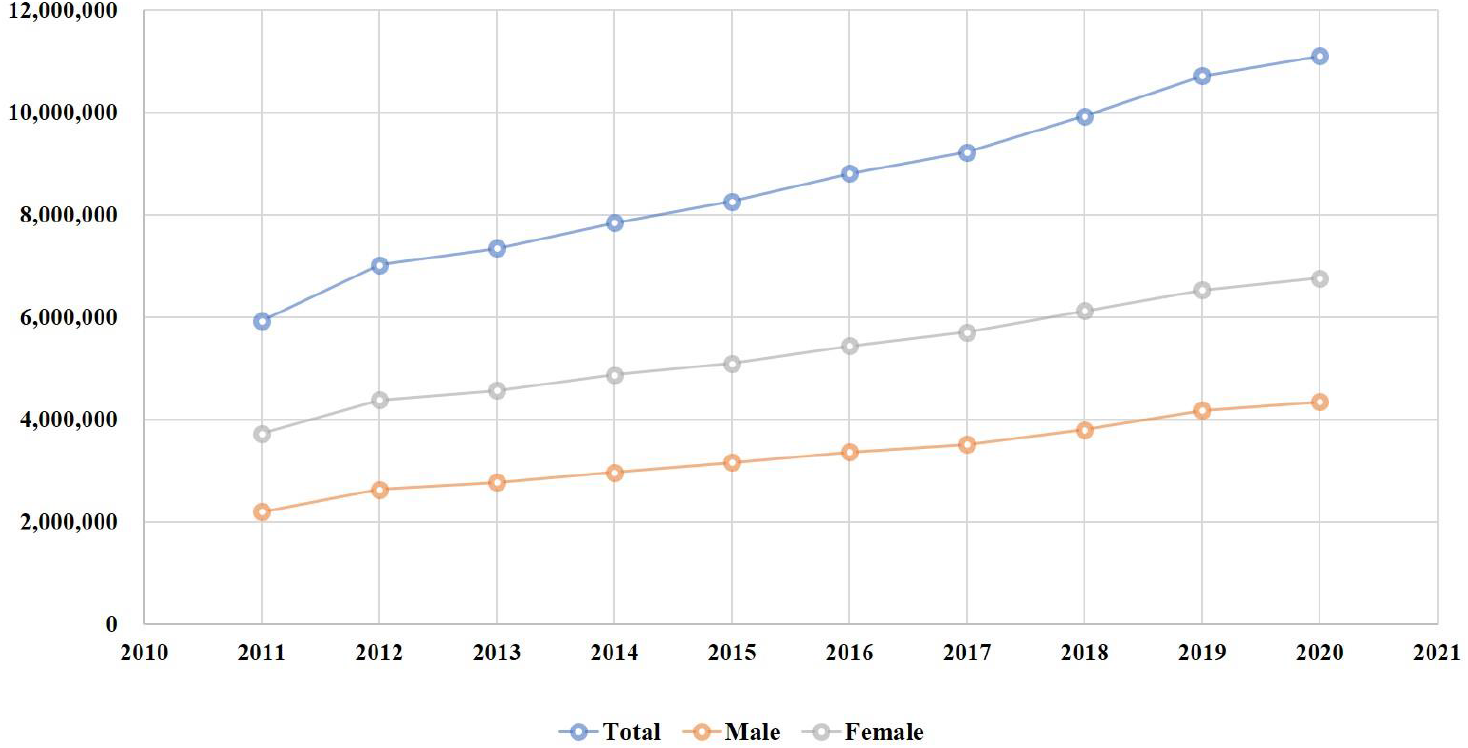
Annual use of medical services for sleep disorders by sex (2011–2020)

**Table 2.** Use of medical services by sex

| Variables | Total | Male | Female | p-value |
| --- | --- | --- | --- | --- |
| Total number of medical visits | 86,242,369 | 32,983,344 | 53,259,025 |  |
| Number of medical visits per patient | 11.5 (± 26.6) | 10.9 (± 25.4) | 11.7 (± 27.4) | <0.0001 |
| Type of medical service |  |  |  |  |
| Korean medicine | 8,323,747 (9.7%) | 2,257,536 (6.8%) | 6,066,211 (11.4%) | <0.0001 |
| Western medicine | 77,918,622 (90.3%) | 30,725,808 (93.2%) | 47,192,814 (88.6%) |  |
| Type of medical visit |  |  |  |  |
| Inpatient admission | 2,488,588 (2.9%) | 1,478,311 (4.5%) | 1,010,277 (1.9%) | <0.0001 |
| Outpatient visit | 83,753,781 (97.1%) | 31,505,033 (95.5%) | 52,248,748 (98.1%) |  |
| Medical expenses per visit |  |  |  |  |
| Total medical expenses | 71,931 (± 372,377) | 91,654 (± 453,808) | 59,716 (± 310,819) | <0.0001 |
| Patient charge | 12,785 (± 56,287) | 14,697 (± 63,616) | 11,601 (± 51,188) | <0.0001 |
| Drug prescription fee | 11,629 (± 161,062) | 12,651 (± 244,311) | 10,996 (± 71,006) | <0.0001 |
| Medical expenses per patient |  |  |  |  |
| Total medical expenses | 830,707 (± 4,595,063) | 1,002,640 (± 5,476,533) | 714,282 (± 3,881,934) | <0.0001 |
| Patient charge | 147,655 (714,887) | 160,782 (± 792,815) | 138,765 (± 138,765) | <0.0001 |
| Medication prescription fee | 134,300 (± 929,518) | 138,394 (± 1,107,515) | 131,528 (± 786,423) | <0.0001 |

Data are represented as number and proportion (%) or mean ± standard deviation.

### Medication prescriptions and others

Over the past decade, medications covered by the National Health Insurance for Sleep Disorders in Korea included 51,095,781 (47.7%) prescriptions for benzodiazepines, 31,325,590 (29.2%) prescriptions for non-benzodiazepines, and 17,457,246 (16.3%) prescriptions for antidepressants. As for the National Health Insurance, the herbal medicines prescribed for sleep disorders were 100,065 cases of Gamisoyo-san, 23,841 cases of Hwanglyeonhaedok-tang, and 11,490 cases of Sosiho-tang (Figure 4). Excluding herbal medicines, acupuncture was the most frequently prescribed treatment (13,727,099 cases), followed by moxibustion, hot/cold physical therapy, electronic needle stimulation, and cupping (Supplementary Figure 1).

**Figure 4.**
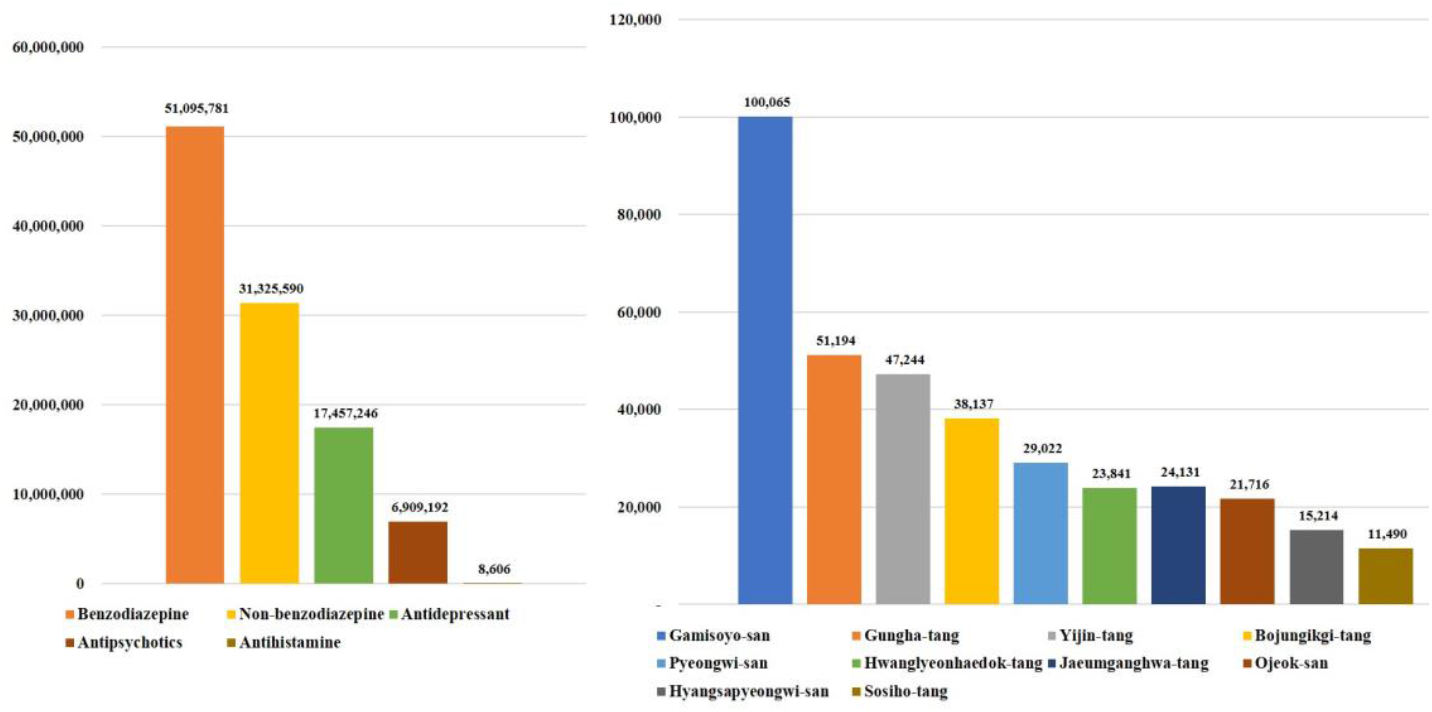
Medication prescriptions for sleep disorders by medical type.

## DISCUSSION

The prevalence of sleep disorders in Korea has nearly doubled over the past 10 years, from 9.4% in 2011 to 17.3% in 2020. Similarly, sleep disorder prevalence in countries like the United States, Norway, England, Germany, and Japan has also shown an overall increasing trend^7-9,11-13,15,17^. Several possible explanations exist for this recent increase in the prevalence of sleep disorders. First, it can be assumed that primary or secondary sleep disorders will increase in absolute numbers, and secondary sleep disorders, such as insomnia, are caused by the use of drugs and substances or by medical or psychiatric diseases and are known to account for more than 70% of all insomnia^31^. The prevalence of secondary sleep disorders also increases with the prevalence of cancer, Parkinson’s disease, and degenerative cerebrovascular diseases such as dementia and obesity, which are well-known causes of sleep disorders^32-34^. Furthermore, since 2020, the number of people complaining about sleep disorders has increased worldwide, which could be the aftermath of the COVID-19 pandemic^35^. The second reason may be the behavioural change in seeking medical services to solve the problem, as patients’ perceptions of sleep disorders change. Previously, sleep disorders, such as insomnia, were regarded as secondary symptoms resulting from diseases rather than a single disease. However, as ICSD-2 was revised to ICSD-3; hence, insomnia disorder was suggested^24^. Accordingly, there has been an apparent change in insomnia recognition as a disease requiring treatment, such as paying attention to classification according to Objective Sleep Duration (OSD) in clinical practice and applying it to treating insomnia cases^36^. In 2017, the American Academy of Sleep Medicine and European Academy of Sleep Medicine published guidelines for insomnia treatment that preferentially recommend cognitive behavioral therapy (CBT-I)^23,37^.

Hospital visits owing to sleep disorders were more frequent among women than among men. Previous studies have shown sex differences in the prevalence and types of sleep disorders. A meta-analysis of sex differences in the prevalence of insomnia reported a significantly higher prevalence in women^38^. The high prevalence of sleep disorders in women, including insomnia, is multifactorial. It may be because women are more likely to experience physical problems such as osteoporosis, fractures, and joint diseases, which affect sleep quality, and they have a higher risk of developing psychiatric problems such as depression and anxiety, resulting to increase insomnia risk^39^. According to the results of this study, insomnia accounts for more than 90% of all sleep disorders. Hence, the above factors can explain women’s higher prevalence of sleep disorders. In contrast, in the case of obstructive sleep apnea (OSA), the proportion of all sleep disorders is low, so it does not have a significant effect on the overall prevalence; however, it is well-known that it has a higher prevalence in men^40,41^. The current study confirmed that the average age for sleep disorder diagnosis in Korea is 53.5 years old and that the number of patients with sleep disorders tends to increase. There are many elderly patients with sleep disorders because sleep quality decreases with age owing to the apparent evolutionary biological relationship between aging and sleep. Many sleep-related studies have reported that subjective and objective sleep quality deteriorates with age^42,43^. Moreover, according to a study comparing the sleep of older and younger adults, older adults spend less time in slow-wave sleep, resulting in a reduced percentage of deep sleep^44^. As individuals age, sleep becomes more fragmented, leading to changes in sleep stages and more frequent awakenings^45^. Sleep homeostasis decreases with age^46^. Furthermore, the prevalence of diseases that cause sleep disorders, such as metabolic disorders, cardiovascular diseases, neurodegenerative diseases, and cancer, increases with age^47-51^.

Insomnia is the most common sleep disorder among the sleep disorder types, according to ICSD-3, and approximately 17.5% of Koreans visit hospitals for insomnia. Insomnia is “a persistent difficulty with sleep initiation, duration, consolidation, or quality that occurs despite adequate opportunity and circumstances for sleep and results in some form of daytime impairment”^52^. According to another study (2005 – 2013) that reported the prevalence and incidence of insomnia in Korea, 5.78% of adults aged 20 years or older had insomnia^53^. Additionally, approximately 17% of the total subjects in a study reported insomnia in 2002, and 22.8% of subjects in a 2009 study reported insomnia^16,17^. Insomnia is Korea’s most common sleep disorder, and its incidence steadily increases.

Over the past 10 years, the mean number of hospital visits due to sleep disorders in Korea was 11.5, and the mean total medical expense for one patient during the observation period was 830,707 KRW (629.13 USD). The insomnia related examination cost $3,508,635 which was about half of total insomnia-related medical expenses^54^. Australia reported total direct health expenditures related to inadequate sleep of approximately 1.24 billion AUD from 2016–2017^55^. In the United States, the direct medical cost due to insomnia in 1995 was estimated at approximately $13.9 billon^56^, and in a review paper on the health economics of insomnia treatment published in 2016, it exceeds $100 billion per year for insomnia treatment^57^. In the United States, direct/indirect social costs due to insomnia increased approximately 10 times over nearly 20 years, most of which contributed to indirect costs such as reduced work performance, increased use of medical care, and increased risk of accidents^57^. The social cost to be paid due to sleep disorders is increasing at a non-negligible rate over time, and improving sleep quality seems to be a social task to be solved.

Among the prescription drugs for sleep disorders, benzodiazepines accounted for 47.8%, and non-benzodiazepines accounted for 29.3%. According to the Korean Neuropsychiatric Association guidelines for insomnia treatment, CBT is recommended first^58^. Sleeping pills are an effective and easy method for treating insomnia, but they carry the risk of dependence and abuse. The KFDA and treatment guidelines recommend shortening drug treatment for unremedied insomnia, mostly within four weeks. In Korea, approximately 10% of patients were prescribed herbal medicine for treating sleep disorders. In practice, medicines in Western medicine and herbal medicines in Korean medicine are prescribed separately or together to treat sleep disorders. However, studies on the effects or side effects of combined prescribed and herbal medicines in patients with sleep disorders are lacking.

The data source of the present study has the advantage of being able to estimate the medical use behaviour of the entire population but also has the limitation of being able to confirm only the items subject to health insurance benefits and review. Information on uncovered herbal medicines or decoctions not subject to health insurance review could not be counted for the herbal medicine. Additionally, the results of tests for the diagnosis of sleep disorders were not confirmed. Nevertheless, we believe it will be meaningful because we have confirmed the current address of the prevalence and treatment of sleep disorders in the last 10 years.

In conclusion, sleep disorders are continuously increasing with more individuals seeking medical intervention resulting in increased personal and social medical expenses. Sleep disorders should be recognized as a significant health problem that needs to be actively addressed to improve quality of life.

## Data Availability

The data that support the findings of this study are available from the Healthcare Big Data Hub of the HIRA. However, restrictions apply with regard to availability as they were used under license for research in the current study; therefore, these data are not publicly available.

## Contributors

Conceptualization was done by EKA, YB, and HJJ. Methodology was done by EKA. The original draft was written by EKA and HJJ. Review and editing were done by YB, JP, SL and HJJ. Funding was acquired by JP and SL. Supervision was done by HJJ.

## Acknowledgements

We would like to thank Editage (www.editage.co.kr) for English language editing.

## Figure legends

Supplementary figure 1. Treatment prescriptions of Korean medicine, excluding herbal medicine

