## Supplementary material for "Elevated prevalence and treatment of sleep disorders from 2011 to 2020; a nationwide population-based retrospective cohort study in Korea": suplementary tables and figure

Supplementary table 1. The third edition of International Classification of Sleep Disorders (ICSD-3) and its diagnostic codes

| Category | KCD-7* | Diagnosis |
|---|---|---|
| 1. Insomnia | F51 | Nonorganic sleep disorders |
| | F51.0 | Nonorganic insomnia |
| | F51.9 | Nonorganic sleep disorder, unspecified |
| | G47 | Sleep disorders |
| | G47.0 | Disorders of initiating and maintaining sleep |
| | G47.9 | Sleep disorder, unspecified |
| 2. Sleep related breathing disorders | G47.3 | Sleep apnea |
| 3. Sleep-related movement disorders | G25.8 | Other specified extrapyramidal and movement disorders (restless legs syndrome) |
| 4. Circadian rhythm sleep-wake disorders | F51.2 | Nonorganic disorder of the sleep-wake schedule |
| | G47.2 | Disorders of the sleep-wake schedule |
| 5. Central disorders of hypersomnolence | G47.4 | Narcolepsy and cataplexy |
| | R40.0 | Somnolence |
| | F51.1 | Nonorganic hypersomnia |
| | G47.1 | Disorders of excessive somnolence |
| 6. Parasomnia | F51.3 | Sleepwalking |
| | F51.4 | Sleep terrors |
| | F51.5 | Nightmares |
| 7. Other sleep disorders | F51.8 | Other nonorganic sleep disorders |
| | G47.8 | Other sleep disorders |

* KCD, Korean Standard Classification of Diseases

Supplementary table 2. Medications for sleep disorders in South Korea

| Category | Drugs with indications for sleep disorders |
|---|---|
| Benzodiazepine | Flurazepam[a], Triazolam[a], Flunitrazepam[a], Brotiazolam[a], Clonazepam |
| Non-benzodiazepine<br>Non-benzodiazepine GABA modular (z-class) | Zolpidem immediate-release[a], Zolpidem controlled-release[a], Eszopiclone[a] |
| Antidepressant | Trazodone, Mirtazapine, Amitriptyline, Doxepin[a] |
| Antihistamine | Doxylamine[a], Diphenhydramine[a] |
| Melatonin | Prolonged-release melatonin[a] |
| Antipsychotics | Quetiapine, Olanzapine |
| Herbal medicines[a] | Gamisoyo-san[b], Hwanglyeonhaedok-tang[b], Galgeunhaegui-tang[b], Dangguiyukhwang-tang[b], Sosiho-tang[b]<br>Gwibi-tang, Sanjoin-tang, Eoggansangajinpibanha, Ongyeng-tang, Chenwangbosim-dan, Gyejigagolmoreo-tang, Ondam-tang, Samhwangsasim-tang, Galgeunhwanglyeonhwanggeum-tang, Sihogyejigeongang-tang, Seungma |

[a] Approved by Korea Food and Drug Administration (KFDA)

[b] Medicines covered by national health insurance benefits in South Korea

Supplementary table 3. List of the 56 herbal medicines which are covered by national health insurance in South Korea

| No. | Romanization of Korean | Chinese | Japanese |
|---|---|---|---|
| 1 | Gamisoyo-san | Jiaweixiaoyao-san | Kamishoyo-san |
| 2 | Galgeun-tang | Gegen-tang | Kakkon-to |
| 3 | Galgeunhaegui-tang | Gegenchengqi-tang | N/A |
| 4 | Gumiganghwal-tang | Jiuweiqianghuo-tang | Kumikyokatsu-to |
| 5 | Gungso-san | Qiongsu-san | N/A |
| 6 | Gungha-tang | Qiongxia-tang | N/A |
| 7 | Naeso-san | Neixiao-san | Naishou-san |
| 8 | Danggwiyeongyo-eum | Dangguilianqiao-yin | N/A |
| 9 | Dangguiyukhwang-tang | Dangguiliuhuang-tang | Tokirikuoto |
| 10 | Daeshiho-tang | Dachaihu-tang | Daisaiko-to |
| 11 | Daecheonglyong-tang | Daqinglong-tang | N/A |
| 12 | Daehwajung-eum | Dahezong-yin | N/A |
| 13 | Daehwangmokdanpi-tang | Dahuangmudan-tang | N/A |
| 14 | Doinseunggi-tang | Taorenchengqi-tang | N/A |
| 15 | Banhabakchulcheonma-tang | Banxiabaizhutianma-tang | Hangebyakujutsutenma-to |
| 16 | Banhasasim-tang | Banxiaxiexin-tang | Hangeshashin-to |
| 17 | Banhahubak-tang | Banxiahoupo-tang | Hange Koboku-To |
| 18 | Baekchool-tang | Baizhu-tang | N/A |
| 19 | Bojungikgi-tang | Buzhongyiqi-tang | Hochuekki-To |
| 20 | Boheo-tang | Buxu-tang | N/A |
| 21 | Bokryongbosim-tang | Fulingbuxin-tang | N/A |
| 22 | Bulhwangeumjeonggi-san | Buhuanjinzhengqi-san | Fukankinshoki-san |
| 23 | Samsoeum | Shensuyin | Jinsoin |
| 24 | Samchulgeonbi-tang | Shenzhujianpi-tang | Sanjutsukenhi-to |
| 25 | Samhojagyak-tang | Shenhushaoyao-tang | N/A |
| 26 | Samhwangsasim-tang | Sanhuangxiexin-tang | N/A |
| 27 | Saengmaek-san | Shengmai-san | Seimyaku-san |
| 28 | Sosiho-tang | Xiaochaihu-tang | Shosaiko-to |
| 29 | Socheongryong-tang | Xiaoqinglong-tang | Shoseiryu-to |
| 30 | Seungyangbowi-tang | Shengyangbuwei-tang | N/A |
| 31 | Sigyeongbanha-tang | Chaigengbanxia-tang | N/A |
| 32 | Sihogyeji-tang | Chaihuguizhi-tang | Saikokeishito |
| 33 | Sihosogan-tang | Chaihushugan-tang | N/A |
| 34 | Sihocheonggan-tang | Chaihuqinggan-tang | N/A |
| 35 | Antae-eum | Antai-yin | N/A |
| 36 | Yeonkyopaedok-san | Lianqiaobaidu-san | N/A |
| 37 | Orim-san | Wulin-san | N/A |
| 38 | Ojeok-san | Wuji-san | Goshaku-san |
| 39 | Yijung-tang | Lizhong-tang | Richu-to |
| 40 | Yijin-tang | Erchen-tang | Nichin-to |
| 41 | Ikwiseungyang-tang | Yiweishengyang-tang | N/A |
| 42 | Insampaedok-san | Renshenbaidu-san | Ninjinhaidoku-san |
| 43 | Injinho-tang | Yinchenhao-tang | Inchin-ko-to |
| 44 | Jaeumganghwa-tang | Ziyinjianghuo-tang | Jiinkoka-to |
| 45 | Jowiseunggi-tang | Tiaoweichengqi-tang | Choi-Joki-To |
| 46 | Cheongsanggyeontong-tang | Qingshangjuantong-tang | Seijokentsuto |
| 47 | Cheongseoikgi-tang | Qingshuyiqi-wan | N/A |
| 48 | Cheongwi-san | Qingwei-san | N/A |
| 49 | Palmul-tang | Bawu-tang | N/A |
| 50 | Pyeongwi-san | Pingwei-san | Heii-san |

| | | | |
|---|---|---|---|
| 51 | Haengso-tang | Xingsu-tang | N/A |
| 52 | Hyangsapyeongwi-san | Xiangshapingwei-san | Koshaheii-san |
| 53 | Hyeonggaeyeongyo-tang | Jingjielianqiao-tang | Keigai-Rengyo-to |
| 54 | Hwanggeumjagyak-tang | Huangqinshaoyao-tang | N/A |
| 55 | Hwanglyeonhaedok-tang | Huanglianjiedu-tang | Orengedoku-to |
| 56 | Hoechunyanggyeok-san | Huichunliangge-san | N/A |

Supplementary table 4. Annual prevalence of sleep disorders for ten years by sex (2011-2020)

| Years | Total Population* | Total patient | Total prevalence (%) |
|---|---|---|---|
| 2011 | 50,734,284 | 3,867,975 | 7.6 |
| 2012 | 50,948,272 | 4,533,300 | 8.9 |
| 2013 | 51,141,463 | 4,829,063 | 9.4 |
| 2014 | 51,327,916 | 5,196,239 | 10.1 |
| 2015 | 51,529,338 | 5,517,411 | 10.7 |
| 2016 | 51,696,216 | 5,900,528 | 11.4 |
| 2017 | 51,778,544 | 6,126,281 | 11.8 |
| 2018 | 51,826,059 | 6,552,777 | 12.6 |
| 2019 | 51,829,023 | 7,076,228 | 13.7 |
| 2020 | 51,667,688 | 7,446,846 | 14.4 |
| Years | The male population* | Patient (male) | Prevalence (male, %) |
| 2011 | 25,406,934 | 1,477,614 | 5.8 |
| 2012 | 25,504,060 | 1,741,659 | 6.8 |
| 2013 | 25,588,336 | 1,868,062 | 7.3 |
| 2014 | 25,669,296 | 2,016,429 | 7.9 |
| 2015 | 25,758,186 | 2,162,156 | 8.4 |
| 2016 | 25,827,594 | 2,311,577 | 9.0 |
| 2017 | 25,855,919 | 2,395,403 | 9.3 |
| 2018 | 25,866,129 | 2,578,451 | 10.0 |
| 2019 | 25,864,816 | 2,842,071 | 11.0 |
| 2020 | 25,841,029 | 2,987,309 | 11.6 |
| Years | The female population* | Patient (female) | Prevalence (female, %) |
| 2011 | 25,327,350 | 2,390,361 | 9.4 |
| 2012 | 25,444,212 | 2,791,641 | 11.0 |
| 2013 | 25,553,127 | 2,961,001 | 11.6 |
| 2014 | 25,658,620 | 3,179,810 | 12.4 |
| 2015 | 25,771,152 | 3,355,255 | 13.0 |
| 2016 | 25,868,622 | 3,588,951 | 13.9 |
| 2017 | 25,922,625 | 3,730,878 | 14.4 |
| 2018 | 25,959,930 | 3,974,326 | 15.3 |
| 2019 | 25,964,207 | 4,234,157 | 16.3 |
| 2020 | 25,826,659 | 4,459,537 | 17.3 |

*The population of each category is based on the population at the end of December each year published by the Ministry of the Interior and Safety's Population Data of Statistical Yearbooks.

Supplementary figure 1. Treatment prescriptions of Korean medicine, excluding herbal medicine

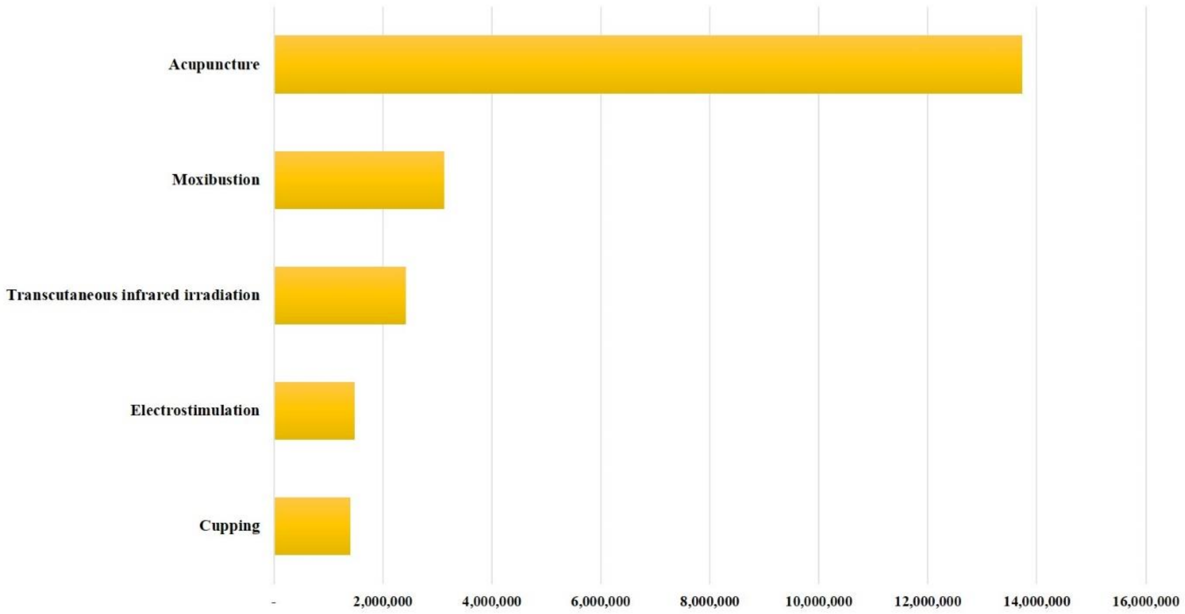
